# Validation of N95 filtering facepiece respirator decontamination methods available at a large university hospital

**DOI:** 10.1101/2020.04.28.20084038

**Authors:** Krista R. Wigginton, Peter J. Arts, Herek Clack, William J Fitzsimmons, Mirko Gamba, Katherine R. Harrison, William LeBar, Adam S. Lauring, Lucinda Li, William W. Roberts, Nicole Rockey, Jania Torreblanca, Carol Young, Loïc G. Anderegg, Amy M. Cohn, John M. Doyle, Cole M. Meisenhelder, Lutgarde Raskin, Nancy G. Love, Keith S. Kaye

## Abstract

**Importance:** Filtering facepiece respirators, including N95 masks, are a critical component of infection prevention in hospitals. Due to unprecedented shortages in N95 respirators, many healthcare systems have explored reprocessing of N95 respirators. Data supporting these approaches are lacking in real hospital settings. In particular, published studies have not yet reported an evaluation of multiple viruses, bacteria, and fungi along with respirator filtration and fit in a single, full-scale study.

**Objective:** We initiated a full-scale study to evaluate different N95 FFR decontamination strategies and their impact on respirator integrity and inactivating multiple microorganisms, with experimental conditions informed by the needs and constraints of the hospital.

**Methods:** We explored several reprocessing methods using new 3M™ 1860 N95 respirators, including dry (<10% relative humidity) and moist (62-66% relative humidity) heat (80-82 °C) in the drying cycle of industrial instrument washers, ethylene oxide (EtO), pulsed xenon UV (UV-PX), hydrogen peroxide gas plasma (HPGP), and vaporous hydrogen peroxide (VHP). Respirator samples were treated and analyzed for biological indicator inactivation using four viruses (MS2, phi6, influenza A virus, murine hepatitis virus), three bacteria (*Escherichia coli, Staphylococcus aureus, Geobacillus stearothermophilus*), and the fungus *Aspergillus niger*. The impact of different application media was also evaluated. In parallel, decontaminated respirators were evaluated for filtration integrity and fit.

**Results:** VHP resulted in ≥2 log_10_ inactivation of all tested biological indicators. The combination of UV-PX + moist heat resulted in ≥2 log_10_ inactivation of all biological indicators except *G. stearothermohphilus*. Greater than 95% filtration efficiency was maintained following 2 (UV-PX + <10% relative humidity heat) or 10 (VHP) cycles of treatment, and proper fit was also preserved. UV-PX + dry heat was insufficient to inactivate all biological indicators. Although very effective at virus decontamination, HPGP resulted in decreased filtration efficiency after 3 cycles, and EtO treatment raised potential toxicity concerns. The observed inactivation of viruses with UV-PX, heat, and hydrogen peroxide treatments varied as a function of which culture media (PBS buffer or DMEM) they were deposited in.

**Conclusions and Relevance:** High levels of biological indicator inactivation were achieved following treatment with either moist heat or VHP. These same treatments did not significantly impact mask filtration or fit. Hospitals have a variety of scalable options to safely reprocess N95 masks. Beyond value in the current Covid-19 pandemic, the broad group of microorganisms and conditions tested make these results relevant in potential future pandemic scenarios.

## Introduction

Filtering facepiece respirators (FFRs) are a critical component of infection prevention in hospitals. They provide protection for healthcare workers against airborne pathogens, such as *Mycobacterium tuberculosis*, Measles virus and, most recently, against severe acute respiratory syndrome coronavirus 2 (SARS-CoV-2), the causative agent of COVID-19. The N95 respirator is the most commonly used among a variety of FFRs and removes at least 95% of airborne particles (1). When caring for patients with COVID-19, the CDC recommends the use of an N95 or better respirator as preferred personal protective equipment (PPE) (2). Reuse of N95 respirators after decontamination has been considered in the past (3) but, until recently, few studies have addressed the effectiveness and feasibility of decontamination at a scale necessary to respond to shortages during a pandemic. Due to unprecedented shortages in N95 respirators in various countries around the world, many healthcare systems have explored, and in many instances have implemented, reprocessing of N95 respirators.

In March 2020, the CDC issued guidance stating that vaporous hydrogen peroxide (VHP), ultraviolet germicidal irradiation (UVGI), and moist heat were “the most promising FFR decontamination methods” (4). On March 28, 2020, the FDA issued an Emergency Use Authorization (EUA) permitting the Battelle Decontamination System, which utilizes VHP, to be authorized for use in decontaminating “compatible N95 respirators.” Although CDC guidance and FDA authorization of decontamination methods were welcomed and necessary, important data are lacking, most notably data pertaining to virucidal and bactericidal efficacy of the different modalities. The current FDA Enforcement Policy for Face Masks and Respirators (5) includes an intended approach for the EUA permitting process that incorporates biological indicators to demonstrate bioburden reductions as follows: ≥ 3 log_10_ inactivation for viruses and ≥ 6 log_10_ inactivation for either mycobacteria or bacterial spores. Notably, antibiotic-resistant bacteria that cause nosocomial infections (such as methicillin-resistant *Staphylococcus aureus* or MRSA) and that are known to exhibit resistance to some of the FFR treatment modalities are not listed in the regulation. Furthermore, FDA recommends demonstrating decontamination effectiveness using multiple virus indicators, including known and available respiratory coronaviruses (e.g., SARS or MERS). To date, most studies have focused on applying single biological indicators to determine contaminant effectiveness of FFRs. SARS-CoV-2 may be transmitted through droplets or bioaerosols (6-9). How well the different media used to deposit microorganisms in decontamination experiments reflect droplets and bioaerosols is not well understood. At this time, the FDA does not recommend a preferred experimental medium.

Fit testing, which is used to verify that a respirator correctly fits the user and is comfortable, has been used to evaluate the practicality of FFR decontamination technologies. Successfully completing a fit test provides assurance that the expected level of protection is provided to respirator wearers by minimizing the total amount of contaminant leakage into the facepiece. Fit testing is a crucial component of healthcare worker safety and should be conducted at least annually for each type of respirator that a user wears. During the COVID-19 pandemic, fit testing was ramped up at many centers to expand the healthcare workforce capable of safely providing care to patients infected with SARS-CoV-2. In addition to fit testing, N95 filtration performance must be maintained with decontaminated respirators. The NIOSH standard requires 95% of particles challenging the mask, roughly sized between 10 and 300 nm, be prevented from passing through.

At University of Michigan Health System (UMHS), we initiated a full-scale study to evaluate different FFR decontamination strategies, with experimental conditions informed by the needs and constraints of the hospital. We sought to determine the degree to which VHP, pulsed xenon UV (UV-PX), and dry versus moist heat decontamination processes: inactivated several biological indicators applied to FFRs; impacted mask filtration efficiency; and altered mask fit. Inactivation experiments included varying combinations of four viruses (the non-enveloped phage MS2, and the enveloped phage Phi6, influenza A virus, and murine hepatitis virus – a coronavirus), three bacteria (*Escherichia coli, Staphylococcus aureus, Geobacillus stearothermophilus*), and one fungus (*Aspergillus niger*), as well as three different culture and application media. In addition, other critical issues pertaining to FFR reprocessing were studied, including the impact of decontamination methods on respirator filtration and on fit testing results for the wearer. Finally, the issues of practicality and scalability of each reprocessing method were evaluated.

## Method and Materials

We explored several FFR treatment options, including dry and moist heat, ethylene oxide (EtO), UV-PX, hydrogen peroxide gas plasma (HPGP), and vaporous hydrogen peroxide (VHP). The 3M 1860 respirator is the primary N95 FFR used at UMHS; therefore, it was the only type tested with biological indicators and is the primary focus of this paper. Integrity tests for several additional respirator types were conducted, including 3M 8210, 3M 8511, Moldex 1511 and several KN95 masks (Supplement eTable 1). For each method, respirators, respirator sections (e.g., sectioned in half), or coupons cut from respirators were treated and analyzed for biological inactivation, filtration integrity, or fit.

### FFR integrity testing

New respirators, as well as new respirators that had been subjected to different decontamination treatments were assessed for their particle filtration efficiency using a custom-built experimental apparatus (Supplement eFigure 1). Assessments were based on differences in particle penetration and differential pressure across each mask as a function of processing method and number of decontamination cycles (Supplement eTable 2). Details of the FFR integrity testing are provided in the Supplement. Briefly, a protocol for evaluating respirator filtration efficiency was developed based on NIOSH standard TEB-APR-STP-0059. FFRs were mounted across a small duct through which NaCl aerosols flowed. Size-resolved measurements of aerosol electrical mobility diameter and condensation mediated particle counts upstream and downstream of the FFR were used to determine aerosol penetration, the complement of mask filtration efficiency. Our protocol deviates in several ways from the NIOSH standard, which requires pre-conditioning masks for 24 hours and challenging masks until they reach a particle loading of 200 mg. Nevertheless, our protocol maintains much of the NIOSH standard and can be used to qualitatively differentiate the relative effects of different decontamination treatments, or repeated treatment on FFR integrity as indicated by filtration efficiency and pressure drop.

In addition, fit testing was performed by the Occupational Health Services (OHS) in the hospital with decontaminated respirators according to hospital protocols, which follow OSHA guidelines (10). This process includes the completion of a medical surveillance questionnaire to ensure the individual to be tested meets required criteria. Upon Provider clearance, the individual to be fit tested is put through a “taste test” with saccharin and/or bitrex and is subjected to a series of other tests and movements. An FFR passes the fit test when it is found to be appropriately sealed to the wearer’s face with no leaks.

### Decontamination

#### Heat

Due to the need to scale treatment for up to 3,000 N95 respirators per day, we explored the possibility of using the dry cycle of industrial instrument washers available at UMHS for decontamination. The dry cycle essentially simulates an oven with forced air convection. The temperature range of these washers is 82 - 93 °C with a 30-minute cycle time. When the temperature is set to 82 °C for 30 minutes, there are approximately 10 minutes of warm up in the 30-minute cycle, resulting in 20 minutes at 82 °C (Supplement eFigure 2). Humidity in the washer was varied to achieve three levels of relative humidity (RH = dry, low, or moderate) and was measured in one of two ways. A Fisherbrand Certified Traceable Digital Hygrometer/Thermometer (Basic Model 11-661-7A) was used to measure RH under baseline conditions (*dry*, operating the instrument washer without modification), and with moisture added (*low RH*, seven 43 cm × 66 cm surgical towels receiving 700 mL deionized water). As it was difficult to increase the RH greater than 8-10% (Supplement eFigure 2), we used a method that allows for temperature monitoring and humidity control coupled with mask isolation, as developed by Anderegg et al (11). This method achieves *moderate RH* using Ziploc medium square (rigid) polypropylene sealed containers. Multiple individual containers were placed in the oven, including two containers that contained temperature and humidity sensors (SEK-SCC30-DB-Sensor) to log the environmental conditions over the course of a cycle (Supplement eFigure 3). These sensors were mounted through the lids of the containers with a wired connection that extended out the gasketed washer door to a data acquisition computer (SEK-Sensorbridge). For our experiments, we added ∼300 µL deionized water to a 7.5 × 7.5 cm^2^ paper towel before placing the sealing lids on the containers. For moderate RH heat experiments, containers with sensors and a mask represented the conditions within all sealed Ziploc containers in the instrument washer, and were present along-side containers containing skeleton masks to which experimental coupons were attached (described below).

#### Pulsed Xenon UV (UV-PX)

A LightStrike™ Pulsed Xenon UV lamp (Model PXUV4D, Xenex) was used to deliver polychromatic (200 - 315 nm) wavelengths of UV light across the UV-C and UV-B range. A 4.3 m × 4.3 m UV room was constructed and its walls were prepared with reflective material to maximize UV light coverage throughout the room. N95 respirators or respirator coupons were clipped to wire racks using metal binder clips strung on wire attached to a metal framed rack (clothesline style), keeping the respirators from overlapping. The racks of masks, were positioned around the Xenex robot to maximize the degree to which the UV light contacted the surfaces of all the masks and avoid shadows. Decontamination cycles were 5 minutes, in accordance with recommendations by 3M (12), the FFR manufacturer. The total dosage of the unit was measured across the UV-C wavelengths using a flame irradiance spectrometer (Ocean Insight) and analyzed using Ocean View 2.0 software. Dosage (µW/cm^2^) is determined across wavelength by integrating the area under a plot of μW/cm^2^-nm versus wavelength (nm) (Supplement eTable 3). Fluence (mJ/cm^2^) is calculated for a given distance from the light source based on time of exposure.

#### Ethylene oxide (EtO)

A 3M Steri-Vac 5XL Ethylene Oxide (EtO) Sterilizer/Aerators was evaluated for respirator decontamination. EtO units are low temperature sterilizers that use EtO as its sterilant and have built in aerators, which automatically activate after the sterilization phase is complete. This 3M 5XL EtO sterilizer utilizes one 100% EO cartridge (model number 4-100), per load, to deliver the sterilant into the EtO chamber during the sterilization phase of the cycle. Respirators were placed in a paper/film sterilization pouch, with an EtO chemical indicator-1 mask per sterilization pouch. Pouched masks and an EtO biological indicator vial (*Bacillus atrophaeus* spores) were placed into the EtO chamber and treated with 1-hour exposure at 55 °C, 45% RH at EtO injection, and 12 hrs of aeration. Total cycle time was 15 hours. Sterilization parameters passed and the biological indicator was negative, indicating complete kill of the spores.

#### Hydrogen Peroxide Gas Plasma (HPGP)

A low temperature Sterrad 100NX system was used to treat mask samples with HPGP. Sterrad 100NX units utilize disposable cassettes that contain 59% nominal hydrogen peroxide solution in plastic cell packs. The full decontamination cycle consists of two identical half decontamination cycles. Initially, hydrogen peroxide vapor was introduced into the chamber to allow contact with all items being treated, then electrical energy was added to turn the vapor into gas plasma. After a designated treatment time, the chamber was vented to allow it to return to atmospheric pressure. To limit the risk of cross contamination between dirty and clean, the respirators were packaged in the decontamination room, with staff handling and packaging them while wearing PPE. The N95 respirators or respirator coupons were packaged individually in sealed Tyvek sterilization pouches with chemical indicators and transported to the clean area. The packaged N95 respirators were loaded into the sterilizer chamber with a biological indicator vial containing *Geobacillus stearothermophilus* spores. The sterilization “express cycle” was used for N95 respirator decontamination, which lasted for 24 minutes. Following treatment, sterile packs containing samples were removed from the unit, opened, and respirators were allowed to degas for one hour. Biological indicators always indicated complete spore inactivation.

#### Hydrogen Peroxide Vapor (VHP)

A Bioquell Q10 whole room decontamination system was used to administer vaporized hydrogen peroxide. N95 respirators or respirator coupons were clipped to wire racks using metal binder clips strung on wire attached to a metal framed rack. Treatment consisted of three phases, including an initial phase in which hydrogen peroxide was vaporized and emitted into the room (Gassing), a dwell phase in which hydrogen peroxide levels were maintained (Dwell) and a degassing phase in which hydrogen peroxide was filtered from the air (Aeration). This process was tested twice under two different operating conditions. For Condition 1, Parametric Cycle, settings are automatically determined by the room volume, and settings were as follows: room size set for 105.6 m^3^; Gassing 1 peak reached 446 ppm; Gassing 2 peak reached 495 ppm; Dwell peak reached 490 ppm for 20 minutes; Aeration lasted 1 hour and 8 mins. Total grams of VHP used in Condition 1 were 1,319. For Condition 2, we mimicked the FDA EUA certified method (13) and utilized a timed cycle vs a parametric cycle and increased the amount of hydrogen peroxide and dwell time. With Condition 2, the settings were as follows: Gassing, 135 min at 10 grams per minute, peak reached 659 ppm; Dwell, 150 min run @ 5 grams per min, peak reached 647 ppm; Aeration set to a minimum of 80 mins, with 3 Bioquell aerators to reach 4 ppm to indicate when the cycle was complete. The total amount of VHP injected and used was 2,236 grams. For both runs, mask samples were removed from the room once ambient hydrogen peroxide concentrations fell below 1 ppm, as measured with a Dräger X-am 5100 monitor set to detect hydrogen peroxide. Monitoring was conducted by moving the wire racks of masks into a separate clean room with ambient air. One mask was removed immediately and placed in a closed container with the Draeger sensor and measured for 10 mins. In our experiments, the monitor indicated 0.0 ppm for the duration of the 10 min monitoring period, indicating zero residual VHP on the mask.

### Inactivation experiments

#### Biological Indicators

We used a total of four model viruses (Table 1), three bacteria, and one fungus to study N95 respirator decontamination methods. The viruses included +ssRNA bacteriophage MS2 and dsRNA bacteriophage Phi6 for several reasons, including: they are common human virus indicators in the literature (14, 15); they can be produced at high concentrations that allow for large experimental dynamic ranges; they have rapid turn-around times for culture-based enumeration (12-18 hours), and; they do not require BLS2 or BSL3 facilities. We also employed a recombinant Influenza H3N2 strain because, like SARS-CoV-2, it is an enveloped ssRNA virus that is transmitted via large respiratory droplets, and perhaps small particle aerosols (16). It has surface proteins that are required for infectivity, and influenza viruses were used in previous studies on FFR decontamination (3, 17). The recombinant H3N2 viruses used in this study make firefly luciferase in infected cells, which provides a rapid read-out for infectivity after only 16-18 hours. We also conducted select experiments with the mouse coronavirus murine hepatitis virus (MHV). This virus is in the same genus as SARS-CoV-2 and is therefore expected to behave similarly to it outside of the host. The reason we conducted limited experiments with this virus was due to the limited dynamic range of the assay (1-2 log_10_), and because results took approximately 48 hours to obtain after the completion of experiments. For bacterial inactivation experiments, we used *Escherichia coli* (ATCC 25922), *Staphylococcus aureus* (ATCC 29213), and *Geobacillus stereothermophilus* (ATCC 12980) in different combinations across treatment experiments. *E. coli* was selected to represent a typical Gram-negative hospital pathogen. *S. aureus* is Gram positive and was selected because of its importance as an invasive hospital pathogen that spreads clonally from patient to patient in the hospital, often via the hands of healthcare workers. *G. stearothermophilus* is a thermophilic spore former used to represent infectious agents such as *Clostridium difficile*. Finally, *Aspergillus niger* (a patient isolate) was used to evaluate selected decontamination treatments for fungus inactivation.

**Table 1.** Characteristics of viruses used in this study and SARS-CoV-2.

| Virus | Genome type | Genome size | Particle size (nm) | Enveloped/ Nonenveloped |
| --- | --- | --- | --- | --- |
| SARS-CoV-2 | (+)ssRNA | 29.9 kb | ~100 | Enveloped |
| MS2 | (+)ssRNA | 3.6 kb | ~25 | Nonenveloped |
| φ6 | dsRNA | 13.5 kbp | ~85 | Enveloped |
| Influenza H3N2 | (-)ssRNA | 13.6 kb | ~100 | Enveloped |
| MHV | (+)ssRNA | 31.3 kb | ~120 | Enveloped |

Bacteriophage MS2 and its corresponding *E. coli* host were purchased from American Type Culture Collection (ATCC 15597). MS2 stocks were propagated, enriched, and enumerated based on previously published methods (18). The stocks were filter sterilized with 0.22 µm polyethersulfone (PES) membrane filters. The final MS2 virus stock (∼ 10^11^ PFU/mL) was stored in virus dilution buffer (VDB; 5 mM NaH_2_PO_4_, 10 mM NaCl, pH 7.5) at 4 °C.

Pseudomonas virus phi6 and its host *Pseudomonas syringae* pv. phaseolicola were provided by the Marr laboratory at Virginia Tech. Phi6 was propagated by adding the virus to *Pseudomonas syringae* at 26 °C with a multiplicity of infection (MOI) of 2, followed by incubation for 7 to 9 hours. Cell lysates were filtered through a 0.22 μm PES membrane, and then concentrated with a lab-scale tangential flow filtration system (Millipore, Burlington, MA) with a 30 kDa cellulose filter. The virus concentrate was purified in a 10-40% (wt/wt) step sucrose gradient for 1.5 hours at 120,000 × g at 4 °C, then in a 40-60% (wt/wt) linear sucrose gradient at 120,000 × g for 15 hours at 4 °C. The phi6 virus band was removed and the virus was buffer exchanged into VDB. The final phi6 virus stocks (∼10^12^ PFU/mL) were filtered sterilized through 0.22 μm PES membranes, and stored in VDB at −80 °C.

MHV strain A59 and its supporting cell line DBT were provided by the Leibowitz lab at Texas A&M Health Science Center College of Medicine. DBT cells were grown in Dulbecco’s modified Eagle medium (DMEM) with 10% horse serum, 1% L-glutamine, and 1% penicillin/streptomycin. Cells were incubated at 37 °C and 5% CO_2_. MHV stocks were propagated and enumerated in DBT cells. For MHV propagation, MHV was suspended in DMEM with 2% horse serum, 1% L-glutamine, and antibiotics (referred to as DMEM2) and applied to DBT cells. Following incubation for approximately 24 hours, cells and virus were frozen at −80C. After thawing and centrifugation of the propagation suspension at 3,000 × g for 15 min, virus supernatant was separated from cell debris. The MHV stocks containing approximately ∼10^6^ PFU mL^−1^ in DMEM2 were stored at −80 °C until use.

For influenza virus, we used a recombinant virus that expresses the luciferase reporter in infected cells. This virus is a 6+2 reassortant, in which the genomic segments encoding the surface hemagglutinin (HA) and neuraminidase (NA) are derived from A/Wisconsin/67/2005 (H3N2) and the remaining six segments are derived from A/WSN33 (H1N1). In this case, the segment 3 RNA encodes a polymerase acidic (PA) protein that is fused to the NanoLuc reporter (25). Viruses were rescued following transfection of HEK 293T/MDCK-SIAT1 co-cultures and passed once on MDCK-SIAT1 cells at a multiplicity of 0.05 to generate a passage 1 (P1) stock. All viral passages were performed in Influenza Viral Media (DMEM, 25mM HEPES pH 7.2-7.5, 0.1875% Fraction V BSA, 1% penicillin/streptomycin, and 2 µg/mL TPCK-Treated Trypsin (Worthington Biochemical Corporation)). Viral stocks were stored in 5% glycerol in single use aliquots to avoid additional freeze-thaw cycles. Luciferase expressing influenza viruses were titered by endpoint dilution in 96 well plates, using Influenza Titer Media (same as Influenza Viral Media except 1% BSA). At 18 hours post-infection, media were aspirated and replaced with Influenza Titer Media containing 7.5 µM ViviRen Live Cell Substrate (Promega). Light emission was measured using a BioTek Synergy HTX luminometer with the following settings: 3-minute dark adapting hold, Emission-Hole, Optics Position-Top, GAIN 160, Integration Time-1.00 seconds, Read Height-2.24 mm, room temp. A well was considered positive for infection if the RLU were greater than or equal to twice the average background RLU from eight mock-infected wells.

The three bacterial strains and the fungus were all cultured in the hospital’s Clinical Microbiology Laboratory. Stocks of the bacterial strains were grown on tryptic soy 5% sheep blood agar, and *A. niger* was grown on Sabouraud Dextrose Agar. For the VHP, heat and UV-PX treatments, we also exposed a commercial, autoclave control tab (Bioquell I HPV-B) that contains 10^6^ cfu/ml of *Geobacillus* spores to treatment in order to use a commonly employed, spore-forming biological indicator across all treatments (19). Afterward, both treated and control tabs were placed into a tube with tryptic soy broth and incubated in a 56°C heat block for 72 hours. Turbidity (yes or no) was recorded.

#### Deposition on N95 mask coupons

For virus experiments, circular coupons with 1-inch diameter were prepared from 3M 1860 N95 masks and stapled to keep mask layers from separating. The coupons were weighed and each coupon was placed in a petri dish. A total of 50 uL of either influenza, MHV, or a MS2/Phi6 mixture stock in either PBS or DMEM was deposited on each coupon in 2 uL droplets distributed around the area. The coupons were allowed to dry in biosafety cabinets for approximately 1.25 hours and weighed again to confirm that the coupons had returned to their original mass and were thus dry. Coupons were then transported to the hospital for disinfecting treatments. For each decontamination method, each sample used for treatment had a corresponding no-treatment control. No-virus blank masks were also included to identify possible contamination.

For bacteria and fungus experiments, overnight grown cultures were diluted in saline to a final suspension concentration of 1.5 × 10^6^ or 1.5 × 10^7^ cfu/ml. From there, 50 or 100 μL was applied as multiple drops to rectangular coupons cut from 3M 1860 N95 FFRs that measured 1.25 inches long and 0.25 inches wide. Coupons were allowed to dry before treatment. For each decontamination method tested, one coupon was exposed during treatment, one was a blank that received no culture, and one was an untreated control that received the biological indicator. The latter served to identify non-treatment related inactivation, and was the comparative basis for log reduction calculations. For *S. aureus*, the untreated control had high colony counts and could not be quantified with the dilutions used; therefore, we assumed that the colony count on those plates was equal to the smallest likely number (i.e., if a plate is reported as >100 colony forming units, we assume there are 100 colonies). In cases when the treated coupons had too many colonies to count, this, coupled with the assumptions about counts for the untreated controls, yield maximum log reduction values. *S. aureus* was routinely detected on coupons by several treatments.

#### Extraction from N95 mask coupons

After inactivation experiments, coupons were returned to their respective laboratories for virus extraction and infectivity assays. For all viruses, staples were cut from each coupon and the remaining coupon materials were cut into 5-6 small pieces with sterile scissors and tweezers. The pieces were deposited in 1.3 mL of extraction solutions, which consisted of PBS with 1% BSA (MS2 and Phi6), Dulbecco’s modified Eagle’s medium supplemented with 0.1875% BSA, HEPES, and antibiotics (influenza virus), or DMEM2 (MHV). The tubes containing the coupon pieces and extraction solution were vortexed for 1 min at half-speed. Viruses in the extract solutions were then titered with their respective assays. For the bacteria and mold, all coupons were placed into 8 mL of trypticase soy broth (TSB) and vigorously agitated on a multi-tube vortex unit for 10 minutes. Liquid aliquots (1, 10 or ∼60 μL) were plated in duplicate in trypticase soy sheep blood agar for all strains except *A. niger*, which was plated on Sabouraud Dextrose Agar. *S. aureus* and *E. coli* were incubated at 35°C, *G. stereothermophilus* was incubated at 56°C, and *A. niger* was incubated at 30°C. Plates were counted daily and final reported values are from counts at 72 h.

## Results

### FFR Integrity Under Different Decontamination Treatments

Respirator performance and fit (i.e., integrity) for each treatment are presented in Table 2 for new 3M 1860 FFRs after different decontamination treatments were applied. Respirator integrity test results for other FFR brands are given in Supplement eTable 1. Respirator performance is reported in terms of filtration efficiency and pressure drop measured across the mask at the test flow rate. Biological testing only proceeded if a decontamination treatment was deemed safe based on respirator integrity at the end of decontamination cycles. For this reason, autoclaving (which damaged the FFR so that it was not able to be tested for filtration, pressure drop, or fit) was not tested further. Ultimately, the following decontamination treatments passed the mask integrity testing step and were deemed worthy of additional testing with biological indicators: dry or moist heat, pulsed xenon UV, ethylene oxide (EtO), and both hydrogen peroxide treatment conditions. Of note, mask integrity testing has not yet been performed on FFRs following moderate RH heat treatment.

**Table 2.** Filtration Performance and Fit Test Results for 3M 1860 FFRs after Treatment for Decontamination.

| Decontamination Treatment | Minimum filtration efficiency<br>(# of treatment cycles<sup>a</sup>) | Filtration stability index<sup>b</sup> | Pressure drop<br>(mm H<sub>2</sub>O) | Fit test outcome<br># passed/#tested<br>after (# cycles) |
| --- | --- | --- | --- | --- |
| Dry heat only | >95% (10) | 0.3 | 17.5 | nt |
| UV-PX + dry heat | >97.5% (10) | 0.07 | 17.5 | nt |
| UV-PX + low RH heat | >97.5% (2) | 0.95 | 17.5 | 3/3 (5)<br>2/2 (10) |
| Bioquell VHP | >99% (5) | 0.02 | 17.5 | 3/3 (1)<sup>c</sup> |
| HPGP | > 74%(5) | 5 | 17.5 | nt |
| UV-PX + dry heat + Bioquell VHP | >97.5% (4) | 0.23 | 17.5 | nt |
| EtO | > 99%(1) | 0.1 | 17.5 | nt |

<sup>a</sup>Single cycle duration values, by treatment method using the method designated under Materials and Methods.
<sup>b</sup>Filtration stability index is defined as the ratio of the range of measured filtration efficiency values to the maximum number of treatment cycles for a specified decontamination method. Smaller index values represent less impact of decontamination on filtration performance. <sup>c</sup>Testing with VHP continues; table will be updated when results are available.

### Inactivation of Biological Indicators Under Different Decontamination Treatments

UV-PX + dry or moist heat was evaluated first, followed by chemical-based decontamination methods (hydrogen peroxide-based systems and ethylene oxide).

#### UV-PX + Moist and UV-PX + Dry Heat Treatment

We first explored the combination of UV-PX and heat at 82 °C. Low RH heat was only able to achieve a relative humidity of 8-10% (Supplement eFigure 2). For virus removal under both of these treatments, the influenza virus and the mouse coronavirus MHV exceeded the dynamic range of the assays, achieving greater than 3.9 log_10_ and 1.1 log_10_ inactivation, respectively (Figure 1a). The bacteriophage surrogates MS2 and Phi6 achieved lower inactivation, and inactivation varied from sample to sample and from run to run. We note that in these experiments, the influenza and MHV were deposited on masks in their respective DMEM culture media, whereas the MS2 and Phi6 were deposited in PBS (constituents in Supplement eTables 4 and 5). A control experiment was conducted to assess the potential role of the application media when depositing the viruses onto the masks. Results from this control experiment suggested that MS2 inactivation by UV-PX and heat treatment at 82 °C was higher when the virus was deposited in DMEM as opposed to PBS (Supplement eFigure 5; discussed below). To further probe the impact of matrix, we deposited MS2 and Phi6 in both PBS and DMEM in the remaining experiments.

**Figure 1.**
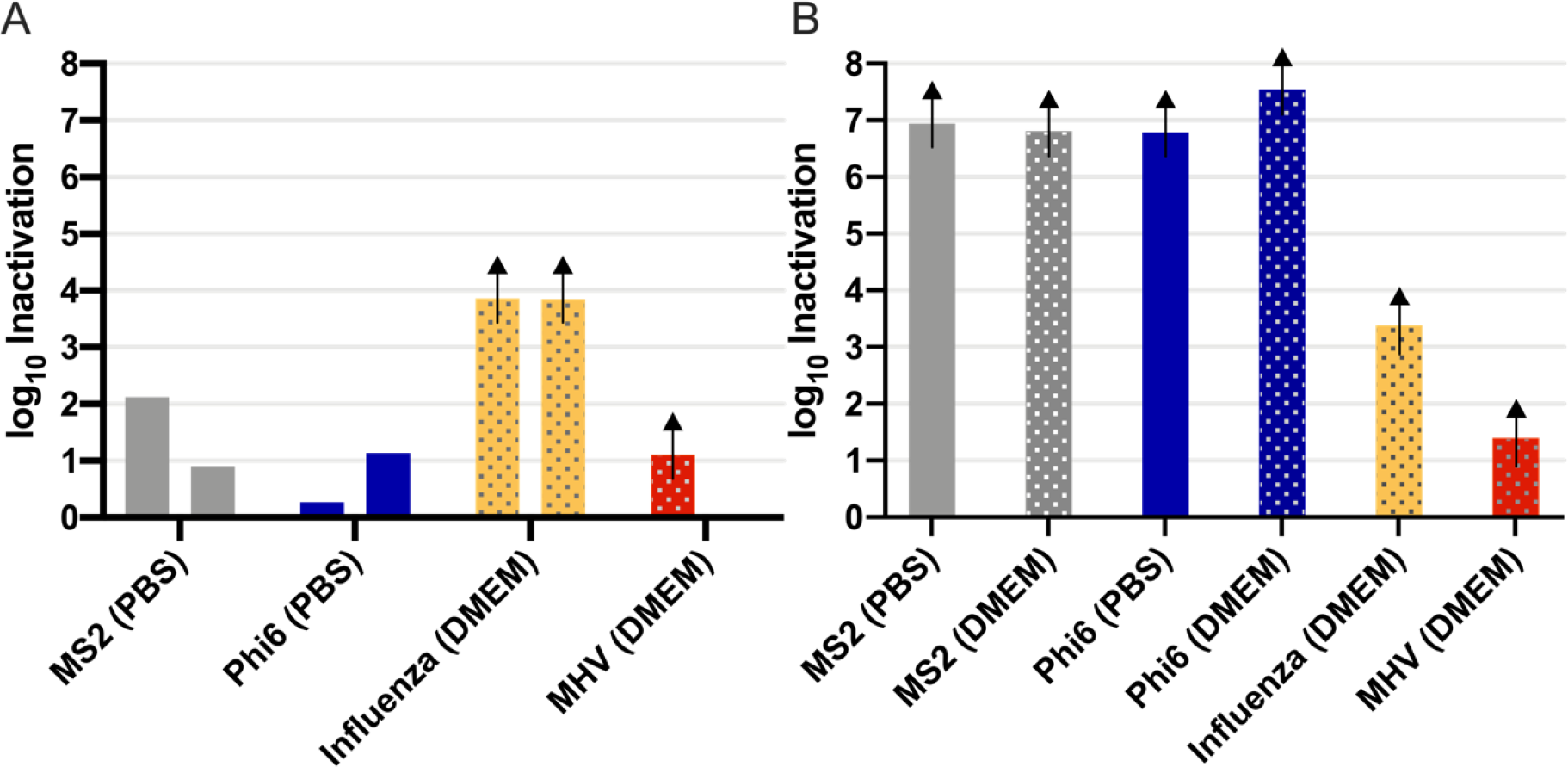
Virus removal from pulsed xenon UV followed by 82 °C heat for 30 mins. (A) Heat treatment with low relative humidity (∼8% RH) and (B) heat treatment with moderate humidity (62-66% RH at 80 °C). Each bar represents the average of replicate experiments conducted on a single day (n = 2-3). Arrows identify samples that exceeded assay detection limits after treatment. Viruses were deposited on the coupons in either PBS or DMEM culture medium.

For the UV-PX + moderate RH + heat experiments with Ziploc containers, temperature reached 70 °C after ∼ 8 mins, then continued to increase while RH decreased until the containers stabilized at 80 °C and 62-66% RH after ∼15 mins, and sustained this condition for ∼ 15 mins (detailed profiles are given in Supplement eFigure 3). The elevated humidity conditions led to increased virus inactivation (Figure 1b). For example, when MS2 was deposited on the coupons in PBS and treated in the oven with ∼8% RH, the observed inactivation was 1-2 log_10_ after a 30-min cycle. In the sealed containers with increased humidity, MS2 inactivation in PBS and DMEM was beyond the assay dynamic range (>6.8 log_10_). Likewise, the inactivation of Phi6 deposited in PBS was less than 1.5 log_10_ when heated to 82 °C and at ∼8% RH, but increased to above the assay dynamic range (>6.6 log_10_) in both PBS and DMEM when treated in sealed containers with moderate humidity. This increased inactivation occurred despite the lower temperatures achieved over 30 minutes, on average, in the container compared to the case when containers were not used and RH never exceeded 10%.

Experiments were also conducted with separate UV-PX and heat treatments. The UV-PX treatment resulted in 0.7 – 1.3 log_10_ MS2, 0.2 – 1.8 log_10_ Phi6, 1.4 – 1.7 log_10_ influenza, and >1.4 log_10_ MHV inactivation (Supplement eFigure 6). The deposition solution appeared to have an impact on the inactivation rate, with viruses deposited in DMEM exhibiting less inactivation on average than the viruses deposited in PBS; however additional experimental replicates need to be conducted to determine statistical differences. When viruses were treated with heat alone at RH>60%, inactivation of all viruses exceeded the dynamic range of the assay (Supplement eFigure 7). Specifically, the MS2, Phi6, influenza virus, and MHV inactivation was >6.8 log_10_, > 6.6 log_10_, > 3.4 log_10_, and >1.4 log_10_, respectively.

We evaluated the inactivation of two bacterial indicators (*S. aureus* and *G. stearothermophilus*; *E. coli* test results will be added) for UV-PX, dry heat, moderate RH heat, and UV-PX with each form of heat (Supplement eTable 6). The *S. aureus* results show that UV-PX and dry heat alone had the highest colony forming unit counts remaining after treatment, with log_10_ reductions <1.0 log_10_. When these treatment steps were used together, inactivation was, at most, 1.2 log_10_. As with the virus results, we saw a large improvement in measured inactivation when humid heat was created with the humidity-controlling Ziploc containers. Inactivation for moderate RH + heat and moderate RH + heat + UV-PX was >2.9 and >2.7, respectively. The spore-forming bacterial indicator *G. stearotherophilus* showed poor inactivation with log reduction levels below 0.3 log_10_ for all treatments. This low level of inactivation was corroborated with commercial sterilization control tabs, which showed positive *Geobacillus* growth under all heat or heat + UV-PX conditions.

#### Hydrogen Peroxide Treatment

We originally tested a Sterrad HPGP system and observed strong virus inactivation (Supplement eFigure 8). Phi6 and influenza virus inactivation exceeded the assay dynamic range following treatment, corresponding to >7.9 and >3.8 log_10_ inactivation, respectively. MS2 was inactivated by an average of 5.6 log_10_. Unfortunately, this system was not scalable to the level necessary for UMHS and mask integrity decreased after just 3 treatments (Table 2). We therefore tested the Bioquell VHP decontamination system under two different conditions (Condition 1-short exposure, and Condition 2-the FDA EUA-approved protocol with long exposure) with all four viruses, including MHV. For Condition 1, we observed >2 log_10_ inactivation for all four viruses (Figure 2). For Condition 2, MS2 and Phi6 were inactivated at >2 log_10_, and MHV showed >1.1 log_10_ inactivation. The results with the VHP treatment highlight that the composition of the deposited virus solution impacts virus inactivation (discussed below).

**Figure 2.**
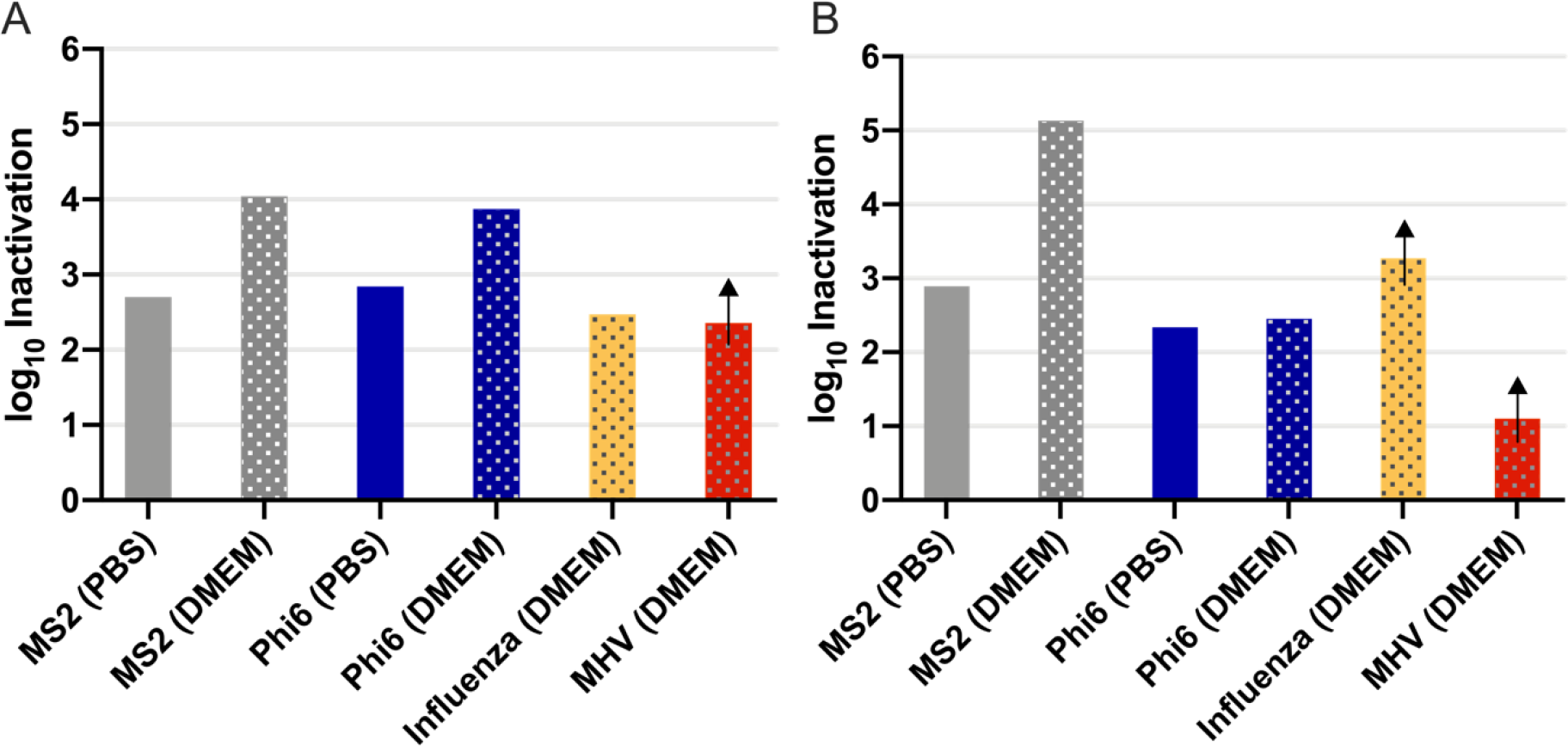
Virus removal from Bioquell VHP system with either A) Condition 1 and B) Condition 2. Each bar represents the average of replicate experiments conducted on a single day (n = 2-3). Solid bars represent experiments where viruses were deposited in PBS and textured bars represent data where viruses were deposited in DMEM. Arrows illustrate samples that exceeded assay detection limits after treatment.

With regards to bacterial results (Supplement eTable 6), the *Geobacillus* spore tab indicator was completely inactivated under both Conditions 1 and 2 as was *E. coli*, which showed no residual colonies on treated coupons (log_10_ inactivation >3.8). In contrast, *S. aureus* was more resistant to inactivation. During Condition 1 when less total hydrogen peroxide was used, only 1 log_10_ inactivation was achieved. This improved to >2.3 and >1.6 log_10_ inactivation with two runs at Condition 2; however, in both runs *S. aureus* colonies were always detected.

#### Ethylene Oxide Treatment

We conducted a single experiment with ethylene oxide and MS2 with triplicate coupons. For each of the three replicates, we exceeded the dynamic range for MS2 assays following treatment (Supplement eFigure 9). This corresponded to a greater than 5.8-log reduction in infective MS2. Although this treatment was effective for inactivating viruses on the masks, we did not expand virus testing beyond this initial experiment nor conduct bacterial experiments, due to concerns about residual EtO and its toxicity to FFR wearers.

#### Impact of deposition solution on virus inactivation

An early control experiment with the sequential UV-PX + low RH heat treatment and MS2 deposited in both PBS and DMEM solutions suggested that the virus application medium may impact inactivation (Supplement eFigure 5). Indeed, follow up experiments with the UV-PX treatment suggested that viruses deposited in DMEM were less susceptible to UV than viruses deposited in PBS (Supplement eFigure 6). This effect was more pronounced for Phi6 inactivation than MS2 inactivation. The average inactivation of Phi6 deposited in PBS was 3.6 log_10_, and only 0.45 log_10_ for Phi6 deposited in DMEM solution. These observations were based on duplicate sample results; additional experiments need to be conducted to characterize the statistical differences between applications with PBS and DMEM. Experiments with moderate RH heat treatment alone with viruses deposited in both PBS and DMEM were inconclusive because in all samples, regardless of the application medium, the viruses were fully inactivated to the assay detection limits (Supplement eFigure 7). Follow up experiments with low RH (10%) and 82°C heat alone conducted in controlled lab experiments emphasize the large impact that media has on heat inactivation (Supplement eFigure 10). For the VHP decontamination method, more inactivation was observed when the MS2 and Phi6 were deposited in DMEM+BSA than when deposited in PBS (Figure 2).

## Discussion

This work was driven by the needs of a major medical center in the midst of the COVID-19 pandemic, and was informed by a multidisciplinary team of investigators. It provides comprehensive information about multiple N95 FFR decontamination methods that were evaluated by assessing the impact of treatment method on mask integrity and ability to inactivate multiple biological indicators. An additional consideration was UMHS’s ability to scale each treatment approach. Using an evaluative process that considered all these factors, a short-list of treatment approaches was selected to assess more deeply with multiple biological indicators under a variety of media matrix conditions. Several important lessons were learned along the way.

Using the FDA’s EUA recommendation to use 3 log_10_ inactivation as a baseline for decontamination of viruses, we found that UV-PX coupled with heat at 80 °C with moderate relative humidity (62-66%) over 15 minutes was sufficient to inactivate all viruses well beyond 3 log_10_ that could be tested to this level. In our sequential UV-PX and heat treatments, most of the observed virus inactivation was achieved by the heat with moderate relative humidity alone, and less than 2 log_10_ was achieved for any virus with the UV-PX treatment alone. Previous work assessing influenza virus removal through moist (85% RH) heat treatment at 65 °C for 30 minutes with viruses applied in a mucin medium demonstrated inactivation to greater than 3 log_10_ (17). With respect to UV, other researchers have reported higher MS2 and influenza log_10_ reductions on N95 respirators following UVC treatment, (3, 17, 20, 21) although the lamps used in these studies generated primarily UV_254_ and provided larger fluence (>1000 mJ/cm^2^) than the Xenex UV-PX unit provided from 200-280 nm at a distance of 1.8 m range (∼24 mJ/cm^2^). To our knowledge, no studies have directly compared inactivation of microorganisms with UVC_254_ to pulsed xenon UVC on N95 respirators.

In contrast to the virus results with heat and UV-PX, the conventional sterilization bacterial spore indicator *Geobacillus* reflected poor inactivation under all heat conditions tested (dry heat, moderate RH heat, and UV-PX); notably, our heat temperature was well below the condition in autoclaves where this indicator is typically used. Furthermore, *S. aureus* achieved >2.9 log_10_ inactivation under UV-PX + humid heat, and UV-PX contributed ≤ 1.0 log_10_ toward this inactivation. The moderate level of *S. aureus* removal by UV-PX is consistent with prior studies in hospital-scale room decontamination studies were conducted (22).

When the Bioquell VHP system was operated according to the FDA EUA-approved condition, we found that inactivation for Phi6 did not exceed 3 log_10_ when applied to masks in multiple media. MS2 only exceeded 3 log_10_ inactivation when in DMEM media, and Influenza exceeded 3 log_10_ inactivation when the Bioquell VHP system was operated according to the FDA EUA-approved condition (Condition 2). Bacterial indicators reveal a potential limitation of VHP. Although both *Geobacillus* and *E. coli* showed complete inactivation, *S. aureus* inactivation was not complete and colonies were always detected on the decontamination coupons. Notably, there is no standard for *S. aureus* inactivation with VHP treatment systems, even though it is known to be catalase-positive and it known to be resistant to VHP treatment (22, 23). Our results suggest that *S. aureus* persistence under VHP may extend to soft fomite surfaces, such as the FFR material evaluated in this study. The implications of this pertaining to hospital infection control needs further consideration, as *S. aureus* (including MRSA) is a major nosocomial threat.

Through this study, we learned that experimental protocols can significantly influence results; specifically, our results demonstrate the impact that virus application medium has on the effectiveness of N95 FFR decontamination. This has practical implications. The FDA EUA recommends treatments achieve >3 log_10_ removal of viruses, but does not give guidance on the application media used to deposit viruses on the masks. The outcomes of this study suggest that >3 log_10_ for a specific virus could depend on the deposition solution. With VHP, for example, the MS2 removal in DMEM was greater than 3 log_10_ under two different treatment conditions, but the MS2 removal in PBS was less than 2 log_10_ under the same conditions. Viruses propagated in tissue cultures are often deposited in the media used for their propagation (e.g., DMEM), which may result in elevated inactivation for heat and VHP. For example, a recently reported non-peer reviewed study (24) conducted with SARS-CoV-2 suggests that 3 log_10_ removal is possible at 70 °C dry heat after 60 minutes; however, the application medium was not defined. If the SARS-CoV-2 was deposited in culture medium and our observed pattern holds, then the SARS-CoV-2 inactivation observed may not be achieved for the virus applied in other media. In contrast to what we observed with heat and VHP, the UV experiments showed that virus deposition in DMEM resulted in less inactivation. This may be due to shielding from constituents present in the complex DMEM solution as compared to PBS (Supplement eTables 4 and 5). Ongoing research should better define the impact of the deposition media constituents on evaluation of decontamination processes, and identify appropriate media conditions that generate conservative estimates of inactivation relative to what occurs in human-derived virus carrier fluids.

Out of the four surrogate viruses, MHV most closely resembles SARS-CoV-2; however, demonstrating 3 log_10_ reduction with this virus was technically challenging due to low MHV stock concentrations and methodologic challenges associated with depositing and recovering viruses from the mask coupons. We could demonstrate at most 1-2 log_10_ inactivation, although we are currently working to increase the dynamic range of the MHV assays to 3 log_10_. The influenza virus experiments had a larger dynamic range, up to 3 to 4 log_10_. The bacteriophage surrogates add additional value to our experiments as they had much higher dynamic ranges, usually >7 log_10_ inactivation and experiments could be carried out quickly and without BSL2 facilities. The combination of four viruses used in this study provides a richer dataset compared to most studies. Influenza virus and MHV are similar in structure to SARS-CoV-2 and therefore provide critical information on how SARS-CoV-2 might behave in the decontamination processes. The two bacteriophages allowed us to quickly develop experimental protocols, probe the impacts of solution composition, and understand the extent of virus inactivation beyond 1-4 log_10_ and for viruses with a range of structures. We are confident that decontamination processes that effectively remove these four viruses will effectively remove SARS-CoV-2. Furthermore, compared to studies that analyze one type of virus in a single medium, these results with a diverse set of viruses in two different media are valuable for evaluation of other viruses of interest in healthcare settings and for potential future pandemic scenarios.

## Conclusions

The COVID-19 pandemic has led to an urgent need for N95 FFR reprocessing. Strategies to decontaminate N95 FFRs should not only inactivate SARS-CoV-2, but ideally, should also inactivate other viral pathogens (such as the influenza virus) as well as bacterial pathogens, particularly those that are multi-drug resistant and can cause outbreaks in the hospital (such as *S. aureus* and *E. coli*). In this manuscript, several N95 FFR decontamination methods were evaluated for their ability to inactivate multiple biological indicators, including viruses and bacteria, while retaining FFR integrity using equipment available at UMHS. Our results suggest that either moist heat (82 °C + 62-66% RH) or VHP can address the hospital’s needs; however, each approach has notable limitations. Moist heat was very effective at eradicating all tested viruses and *S. aureus*, but did not eliminate spore-forming bacteria. Nevertheless, it remains a viable option for decontamination, when coupled with strategic infection control practices (e.g. no reprocessing FFRs that used while caring for *C. difficile* patients). However, a humidified oven is required to reliably achieve the necessary relative humidity to enhance pathogen inactivation. Hydrogen peroxide was effective at inactivating viruses (particularly influenza) beyond 2 log_10_ and often 3 log_10_. However, a notable limitation was our inability to eradicate *S. auerus*. While persistence of *S. aureus* on a reprocessed N95 might impose only limited risk to the wearer, it can serve as a hospital reservoir for this pathogen and facilitate spread to patients. In addition, VHP treatment takes several hours to complete a cycle.

While determining effective methods to decontaminate and reuse N95 FFRs is of paramount importance, it is equally important that methodologic issues that can impact results are noted. For example, experimental limitations, such as the type of culture media used during testing, can change results by orders of magnitude. Better information is needed to understand the characteristics of the carrier matrix that carries the SARS-CoV-2 in a range of environments, and this information needs to be translated into experimental protocols. In addition, while the focus with regards to pathogen eradication needs to be on SARS CoV-2, other pathogens that spread in the hospital and cause significant morbidity should also be considered. There is a need to understand the capabilities and limitations of any N95 FFR decontamination approach to achieve the desired protection against SARS-CoV-2 and to simultaneously achieve the levels of overall infection control desired in hospitals. Ultimately, Eech healthcare setting has different needs, capacity and infrastructure available to address decontamination needs; consequently, we conclude that one solution will not work across all applications.

## Data Availability

Data not presented in the paper or Supplement are available upon request, within the guidelines held by the University of Michigan.

## Acknowledgements

This work was supported by the University of Michigan Health Systems, the University of Michigan College of Engineering, and the Heising-Simons Foundation. Throughout the time this study was underway, our methods were informed by published and pre-published, citable studies with helpful insight from summary reports put forth by N95DECON.org. We would like to acknowledge the contributions of several individuals who contributed to this study in various ways: Andre Boehman, Stephen Ceccio, Jolene Daniel, Bridgette Hegarty, Kathryn Langenfeld, Allen P. Liu, Thomas Mann, Stephanie Prout, Enrique Rodriguez, Charles Solbrig, Drue Stout, Danny Wilson, Margaret Wooldridge. We also want to acknowledge the work conducted by the employees in the University of Michigan Health System Central Sterile Processing Department (CSPD) and all the work conducted by the healthcare workers caring for patients at Michigan Medicine.

## References

1. NIOSH-Approved N95 Particulate Filtering Facepiece Respirators - 3M Suppliers List. Centers for Disease Control and Prevention website. https://www.cdc.gov/niosh/npptl/topics/respirators/disp_part/n95list1.html. Updated April 22, 2020. Accessed April 26, 2020.

2. Infection Control: Severe Acute Respiratory Syndrome Coronavirus 2 (SARS-CoV-2). Centers for Disease Control and Prevention website. https://www.cdc.gov/coronavirus/2019-ncov/hcp/infection-control-recommendations.html. Updated April 13, 2020. Accessed April 26, 2020.

3. Lore MB, Heimbuch BK, Brown TL, Wander JD, Hinrichs SH. Effectiveness of Three Decontamination Treatments against Influenza Virus Applied to Filtering Facepiece Respirators. Ann Occup Hyg. 2012;56(1):92–101. doi:10.1093/annhyg/mer054

4. COVID-19 Decontamination and Reuse of Filtering Facepiece Respirators. Centers for Disease Control and Prevention website. https://www.cdc.gov/coronavirus/2019-ncov/hcp/ppe-strategy/decontamination-reuse-respirators.html. Accessed April 26, 2020.

5. US Food & Drug Administration. Enforcement Policy for Face Masks and Respirators During the Coronavirus Disease (COVID-19) Public Health Emergency (Revised). https://www.fda.gov/regulatory-information/search-fda-guidance-documents. Revised April 2020. Accessed April 26, 2020.

6. Liu Y, Ning Z, Chen Y, Guo M, Liu Y, Gali NK, Sun L, Duan Y, Cai J, Westerdahl D, Liu X, Ho K-F, Kan H, Fu Q, Lan K. Aerodynamic Characteristics and RNA Concentration of SARS-CoV-2 Aerosol in Wuhan Hospitals during COVID-19 Outbreak. bioRxiv. March 2020. doi:10.1101/2020.03.08.982637

7. Chia PY, Coleman KK, Tan YK, Ong SWX, Gum M, Lau SK, Sutjipto S, Lee PH, Son TT, Young BE, Milton DK, Gray GC, Schuster S, Barkham T, De PP, Vasoo S, Chan M, Ang BSP, Tan BH, Leo YS, Ng O-T, Wong MSY, Marimuthu K. Detection of Air and Surface Contamination by Severe Acute Respiratory Syndrome Coronavirus 2 (SARS-CoV-2) in Hospital Rooms of Infected Patients. medRxiv. April 2020. doi:10.1101/2020.03.29.20046557

8. Guo Z-D, Wang Z-Y, Zhang S-F, Li X, Li L, Li C, Cui Y, Fu R-B, Dong Y-Z, Chi X-Y, Zhang M-Y, Liu K, Cao C, Liu B, Zhang K, Gao Y-W, Lu B, Chen W. Aerosol and Surface Distribution of Severe Acute Respiratory Syndrome Coronavirus 2 in Hospital Wards, Wuhan, China, 2020. Emerg Infect Dis. 2020;26(7). doi:10.3201/eid2607.200885

9. Santarpia JL, Rivera DN, Herrera V, Morwitzer MJ, Creager H, Santarpia GW, Crown KK, Brett-Major D, Schnaubelt E, Broadhurst MJ, Lawler JV, Reid P, Lowe JJ. Transmission Potential of SARS-CoV-2 in Viral Shedding Observed at the University of Nebraska Medical Center. medRxiv. March 2020. doi:10.1101/2020.03.23.20039446

10. US Department of Licensing and Regulatory Affairs. Occupational Health Standards. https://www.michigan.gov/documents/CIS_WSH_part451_54075_7.pdf. Amended January 13, 2014. Accessed April 26, 2020.

11. Anderegg L, Meisenhelder C, Ngooi CO, Liao L, Xiao W, Chu S, Cui Y, Doyle JM. A Scalable Method of Applying Heat and Humidity for Decontamination of N95 Respirators During the COVID- 19 Crisis. medRxiv. April 2020. doi:10.1101/2020.04.09.20059758

12. 3M Company. Technical Bulletin: Decontamination Methods for 3M N95 Respirators. Revision 4. https://multimedia.3m.com/mws/media/1824869O/decontamination-methods-for-3m-n95-respirators-technical-bulletin.pdf. Accessed April 26, 2020.

13. Batelle Memorial Institute. Battelle CCDS Critical Care Decontamination System. https://www.battelle.org/docs/default-source/commercial-offerings/industry-solutions/732_battelle-critical-care-decon-system_technical-summaryf.pdf. Accessed April 26, 2020.

14. Rengasamy S, Fisher E, Shaffer RE. Evaluation of the survivability of MS2 viral aerosols deposited on filtering face piece respirator samples incorporating antimicrobial technologies. Am J Infect Control. 2010;38(1):9–17. doi:10.1016/j.ajic.2009.08.006

15. Prussin AJ, Schwake DO, Lin K, Gallagher DL, Buttling L, Marr LC. Survival of the enveloped virus Phi6 in droplets as a function of relative humidity, absolute humidity, and temperature. Appl Environ Microbiol. 2018;84(12). doi:10.1128/AEM.00551-18

16. Yan J, Grantham M, Pantelic J, de Mesquita PJB, Albert B, Liu F, Ehrman S, Milton DK, EMIT Consortium. Infectious Virus in Exhaled Breath of Symptomatic Seasonal Influenza Cases from a College Community. Proc Natl Acad Sci USA. 2018;115(5):1081–1086. doi:10.1073/pnas.1716561115

17. Heimbuch BK, Wallace WH, Kinney K, Lumley AE, Wu C-Y, Woo M-H, Wander JD. A Pandemic Influenza Preparedness Study: Use of Energetic Methods to Decontaminate Filtering Facepiece Respirators Contaminated with H1N1 Aerosols and Droplets. Am J Infect Control. 2011;39(1). doi:10.1016/j.ajic.2010.07.004

18. Pecson BM, Martin LV, Kohn T. Quantitative PCR for determining the infectivity of bacteriophage MS2 upon inactivation by heat, UV-B radiation, and singlet oxygen: Advantages and limitations of an enzymatic treatment to reduce false-positive results. Appl Environ Microbiol. 2009;75(17):5544–5554. doi:10.1128/AEM.00425-09

19. CDC. Sterilization: Sterilization: Monitoring, Division of Oral Health. https://www.cdc.gov/oralhealth/infectioncontrol/faqs/monitoring.html. Accessed April 26, 2020.

20. Fisher EM, Shaffer RE. A Method to Determine the Available UV-C Dose for the Decontamination of Filtering Facepiece Respirators. J Appl Microbiol. 2011;110(1):287–295. doi:10.1111/j.1365-2672.2010.04881.x

21. Vo E, Rengasamy S, Shaffer R. Development of a Test System to Evaluate Procedures for Decontamination of Respirators Containing Viral Droplets. Appl Environ Microbiol. 2009;75(23):7303–7309. doi:10.1128/AEM.00799-09

22. Weber DJ, Rutala WA, Anderson DJ, Chen LF, Sickbert-Bennett EE, Boyce JM. Effectiveness of Ultraviolet Devices and Hydrogen Peroxide Systems for Terminal Room Decontamination: Focus on Clinical Trials. Am J Infect Control. 2016;44(5):e77–e84. doi:10.1016/j.ajic.2015.11.015

23. Pottage T, Macken S, Walker JT, Bennett AM. Meticillin-Resistant Staphylococcus aureus is More Resistant to Vaporized Hydrogen Peroxide than Commercial Geobacillus stearothermophilus Biological Indicators. J Hosp Infect. 2012;80(1):41–45. doi:10.1016/j.jhin.2011.11.001

24. Fischer R, Morris DH, van Doremalen N, Sarchette S, Matson J, Bushmaker T, Yinda CK, Seifert S, Gamble A, Williamson B, Judson S, de Wit E, Lloyd-Smith J, Munster V. Assessment of N95 Respirator Decontamination and Re-Use for SARS-CoV-2. medRxiv. April 2020. doi:10.1101/2020.04.11.20062018

25. Tran V, Moser LA, Poole DS, Mehle A. Highly Sensitive Real-Time In Vivo Imaging of an Influenza Reporter Virus Reveals Dynamics of Replication and Spread. J Virol. 2013;87(24):13321–13329. doi:10.1128/jvi.02381-13

